# Spatial and Temporal Patterns of SARS-CoV-2 transmission in uMgungundlovu, Kwa-Zulu Natal, South Africa

**DOI:** 10.1101/2023.12.08.23299736

**Authors:** Radiya Gangat, Veranyuy Ngah, Rushambwa Tawonga, Justine I. Blanford, Jabulani Ronnie Ncayiyana, Peter Suwirakwenda Nyasulu

**Affiliations:** Division of Epidemiology and Biostatistics, Department of Global Health, Faculty of Medicine and Health Sciences, Stellenbosch University, Cape Town, Western Cape, South Africa; Department of Health, uMgungundlovu Health District Office, Pietermaritzburg, Kwa-Zulu Natal, South Africa; School of Development Studies, University of Kwa-Zulu Natal, Durban, Kwa-Zulu Natal, South Africa; Department of Earth Observation Science (EOS), Faculty of Geo-Information Science and Earth Observation (ITC), University of Twente, Enschede, Netherlands; Division of Public Health Medicine, University of Kwa-Zulu Natal, Durban, Kwa-Zulu Natal, South Africa; Division of Epidemiology and Biostatistics, School of Public Health, Faculty of Health Sciences, University of the Witwatersrand, Johannesburg, South Africa

**Keywords:** SARS-CoV2, Geospatial, Temporal analysis, uMgungundlovu, Kwa-Zulu Natal

## Abstract

**Background:** Investigating the spatial distribution of SARS-CoV-2 at a local level and describing the pattern of disease occurrence can be used as the basis for efficient prevention and control measures. This research project aims to utilize geospatial analysis to understand the distribution patterns of SARS-CoV-2 and its relationship with certain co-existing factors.

**Methods:** Spatial characteristics of SARS-CoV-2 were investigated over the first four waves of transmission using ESRI ArcGISPro v2.0, including Local Indicators of Spatial Association (LISA) with Moran’s “I” as the measure of spatial autocorrelation; and Kernel Density Estimation (KDE). In implementing temporal analysis, time series analysis using the Python Seaborn library was used, with separate modelling carried out for each wave.

**Results:** Statistically significant SARS-CoV-2 incidences were noted across age groups with p-values consistently < 0.001. The central region of the district experienced a higher level of clusters indicated by the LISA (Moran’s I: wave 1 – 0.22, wave 2 – 0.2, wave 3 – 0.11, wave 4 – 0.13) and the KDE (Highest density of cases: wave 1: 25.1-50, wave 2: 101-150, wave 3: 101-150, wave 4: 50.1-100). Temporal analysis showed more fluctuation at the beginning of each wave with less fluctuation in identified cases within the middle to end of each wave.

**Conclusion:** A Geospatial approach of analysing infectious disease transmission is proposed to guide control efforts (e.g., testing/tracing and vaccine rollout) for populations at higher vulnerability. Additionally, the nature and configuration of the social and built environment may be associated with increased transmission. However, locally specific empirical research is required to assess other relevant factors associated with increased transmission.

## Introduction

The SARS-CoV-2 pandemic causing coronavirus disease 2019 (COVID-19) has reached 770, 875 433 confirmed cases worldwide as of 27 September 2023 [4]. Although spatial distribution of infection has been widespread, a distinct asymmetry has been observed in terms of incidence and distribution of cases according to geographical location [3]. With emerging pathogenic outbreaks, it is critical to understand the dynamics of infection transmission. This is of particular importance when analysing the contagion over time and projecting the future epidemiological situation. In this study, Geographic information systems (GIS) were used to analyse the spatial distribution patterns of SARS-CoV-2 in uMgungundlovu district, Kwa-Zulu Natal Province, South Africa, and to highlight the hotspot areas of high transmission.

## Materials and methods

### Study design and participants

A Cross-sectional study design was employed for this study. All laboratory confirmed SARS-CoV-2 positive individuals residing within uMgungundlovu District during each of the four waves experienced to date, formed part of the study population.

### Study setting

The district of uMgungundlovu lies in the interior of Kwa-Zulu Natal (KZN) Province. The district provides health care services to 10% of the KZN population estimated at 1 017 763 [1]. uMgungundlovu covers an area of 9189 square kilometres and is comprised of 7 sub-districts; these include uMsunduzi, Impendle, Richmond, uMngeni, uMkhambathini, uMshwathi and Mpofana (Fig 1). Pietermaritzburg serves as the provincial and legislative capital of the district.

**Fig 1.**
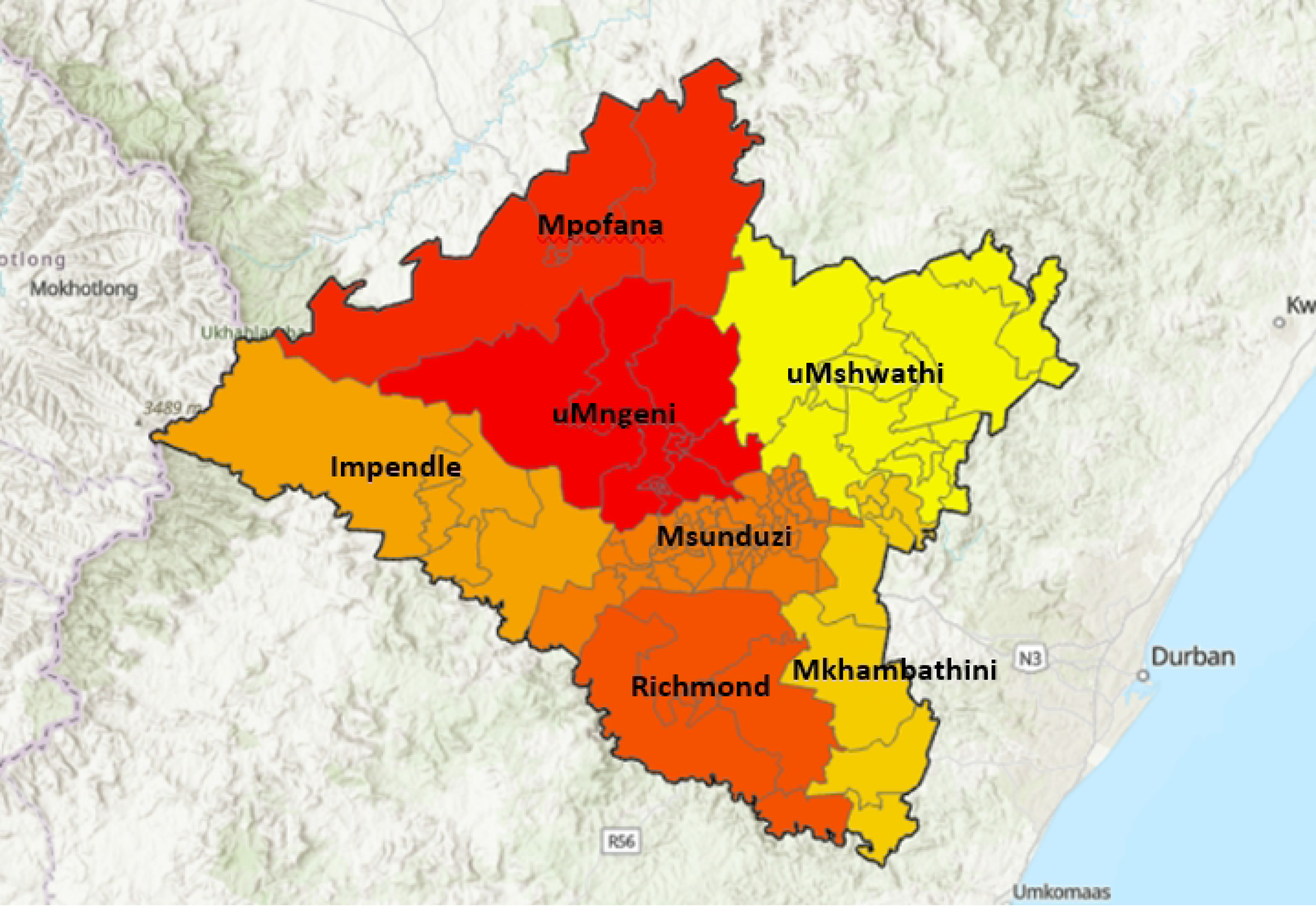
uMgungundlovu sub-district distribution.

### Data collection

During the SARS-CoV-2 pandemic, either nasopharyngeal or oropharyngeal specimens were collected by clinicians throughout the district and sent to laboratories where they were analysed using Polymerase chain reaction (PCR); additionally, rapid antigen testing was conducted at the point of collection. This occurred in the private and public sectors. Data of SARS-COV-2 positive cases are collated daily by the National Institute for Communicable Diseases (NICD) and channelled to the district health office. Data of laboratory-confirmed SARS-CoV-2 cases, including demographic data i.e., age, gender, specimen collection date and ward number were obtained from the NICD daily line lists.

### Data Analysis

Descriptive analysis was conducted separately for each of the four waves. Data were stratified by sex and missing values denoted as NaN. The mean for each independent variable was assessed according to whether the observed differences were statistically significant at the 5% level using the Fisher’s Exact Test, reporting the associated p-value. Statistical analyses were conducted using the R statistical software for data analysis and programming.

In implementing temporal analysis, time series analysis using the Python Seaborn library was used, with separate modelling carried out for each wave. Specimen collection date was used to complete temporal analysis.

Spatial distribution patterns of SARS-CoV-2 were analysed at ward and municipal levels. Base layers for the district used to complete the analysis were obtained from the municipal demarcation board which is an open data source (Table 1). At local municipality level, incidence proportions were analysed using choropleth maps while at ward level, choropleth maps were created using incidence data.

**Table 1.** Base layer data sources.

| *Data* | *Description* | *Source* | *Date*<br>*accessed* | *URL* |
| --- | --- | --- | --- | --- |
| *District*<br>*boundaries* | Arcgis<br>boundary<br>layer | Municipal<br>demarcation<br>board | 23/06/2022 | https://dataportal-mdb-sa.opendata.arcgis.com/maps/97cb14748cef423a94f7b7154387b2dc |
| *Ward and*<br>*municipal*<br>*boundaries* | Arcgis<br>boundary<br>layer | Municipal<br>demarcation<br>board | 23/06/2022 | https://dataportal-mdb-sa.opendata.arcgis.com/maps/97cb14748cef423a94f7b7154387b2dc |

The total number of positive cases were summarized for each ward. The centroid of each ward was obtained and used as input for the KDE analysis.

Moran’s I and LISA (local indicators of spatial association) analyses were used to determine if cases were clustered and if so, to identify where infection clusters or hotspots were located. Cluster analyses were conducted using the Anselin Local Moran’s I statistic to identify potential SARS-CoV-2 hotspots [30]. Positive spatial autocorrelation was found where values at neighbouring locations were similar and referred to respectively as high-high (HH) and low-low (LL). High-high clusters refer to areas of high SARS-CoV-2 incidence occurring near areas of high incidence, and cold spots or low-low clusters occur where areas of low SARS-CoV-2 incidence occur near areas of low incidence. In contrast, negative spatial autocorrelation or spatial outliers occur where dissimilar values occur at neighbouring locations. These are referred to as high-low (HL) and low-high (LH) spatial outliers respectively and occur when areas of high incidence are surrounded by areas of low incidence and vice-versa [5].

A positive value for Moran’s I indicates that a feature has neighbouring features with similarly high or low attribute values (positive spatial autocorrelation) while a negative Moran’s I value indicates that a feature has neighbouring features with dissimilar values; this feature is an outlier. In either instance, the p-value for the feature must be small enough for the cluster or outlier to be considered statistically significant. Statistical significance was set at the 95% confidence level [5]. All analyses were performed using ESRI ArcGISPro v2.0.

### Ethical considerations

Ethics approval was obtained from application [Project ID: 26303, HREC reference number: S22/08/017_COVID-19] submitted to Health Research Ethics Committee (HREC) and was approved via expedited review procedures on 16/09/2022; annual approval was received for 15/09/2023.

Additionally, approval was obtained from the uMgungundlovu District director as well as the Kwa-Zulu Natal provincial department of health.

SARS-CoV-2 incidence data was retrospectively accessed on the 17/09/2022. One author, Radiya Gangat had access to case names in the initial datasets. All case names were replaced with case ID numbers which were in numerical order with no reference to specific individuals before any analysis was completed. Individual case confidentiality was maintained by converting all physical addresses into local municipality names and ward numbers.

## Results

### Description of SARS-CoV-2 transmission

Table 2 shows the number of positive specimens obtained from NICD of SARS-CoV-2 per wave. The highest number of positive specimens were obtained during wave 3.

**Table 2:**
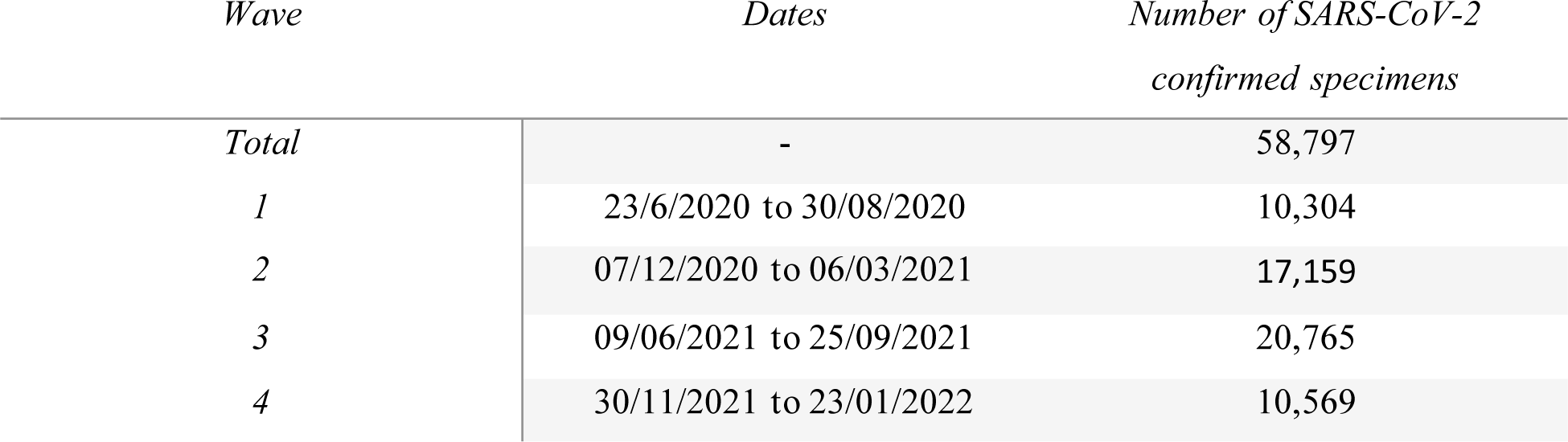
Dates and number of positive specimens per wave.

| *Wave* | *Dates* | *Number of SARS-CoV-2*<br>*confirmed specimens* |
| --- | --- | --- |
| *Total* | - | 58,797 |
| *1* | 23/6/2020 to 30/08/2020 | 10,304 |
| *2* | 07/12/2020 to 06/03/2021 | 17,159 |
| *3* | 09/06/2021 to 25/09/2021 | 20,765 |
| *4* | 30/11/2021 to 23/01/2022 | 10,569 |

During wave 1, 10,304 incident cases were reported as shown in Table 3, 61.7% were females and 38.3% were males. The mean age for males was 40.1 years, with the average slightly higher among females at 40.5 years (p = 0.58). The highest incidence was reported within the age groups 30 to 39 (25%), and the lowest incidence was recorded among patients 80 years and older (1.8%) years (p<0.001).

**Table 3.** Descriptive analysis of SARS-CoV-2 cases, wave 1.

| Grouping<br>Statistics by<br>Gender | N | Overall,<sup>1</sup><br>N=10 304 | Female,<sup>1</sup><br>N=6 360 | Male,<sup>1</sup><br>N=3 944 | P – value<sup>2</sup> | *Incidence*<br>*Proportion*<br>*(per 10 000*<br>*population)* |
| --- | --- | --- | --- | --- | --- | --- |
| Age | 10 287 | 40.34 (17.07)<sup>3</sup> | 40.51 (16.95)<sup>3</sup> | 40.06 (17.24)<sup>3</sup> | 0.58 |  |
| Age Group | 10 286 |  |  |  | <0.001 |  |
| 0-9 years |  | 353/10286 (3.4%) | 181/6350 (2.9%) | 172/3936 (4.4%) |  | 15.1 |
| 10-19 years |  | 708/10286 (6.9%) | 424/6350 (6.7%) | 284/3936 (7.2%) |  | 35.7 |
| 20-29 years |  | 1587/10286 (15.0%) | 1002/6350 (16.0%) | 585/3936 (15.0%) |  | 76.6 |
| 30-39 years |  | 2582/10286 (25.0%) | 1641/6350 (26.0%) | 941/3936 (24.0%) |  | 165.9 |
| 40-49 years |  | 2131/10286 (21.0%) | 1315/6350 (21.0%) | 816/3936 (21.0%) |  | 203.0 |
| 50-59 years |  | 1543/10286 (15.0%) | 947/6350 (15.0%) | 596/3936 (15.0%) |  | 214.1 |
| 60-69 years |  | 835/10286 (8.1%) | 484/6350 (7.6%) | 351/3936 (8.9%) |  | 175.2 |
| 70-79 years |  | 366/10286 (3.6%) | 229/6350 (3.6%) | 137/3936 (3.5%) |  | 202.3 |
| 80+ years |  | 181/10286 (1.8%) | 127/6350 (2.0%) | 54/3936 (1.4%) |  | 282.7 |
| Municipality | 10 304 |  |  |  | 0.96 |  |
| Impendle |  | 96/10304 (0.9%) | 63/6360 (1.0%) | 33/3944 (0.8%) |  |  |
| Mkhambathini |  | 138/10304 (1.3%) | 87/6360 (1.4%) | 51/3944 (1.3%) |  |  |
| Mpofana |  | 204/10304 (2.0%) | 121/6360 (1.9%) | 83/3944 (2.1%) |  |  |
| Richmond |  | 369/10304 (3.6%) | 227/6360 (3.6%) | 142/3944 (3.6%) |  |  |
| Msunduzi |  | 8501/10304 (83.0%) | 5242/6360 (82.0%) | 3259/3944 (83%) |  |  |
| uMngeni |  | 733/10304 (7.1%) | 454/6360 (7.1%) | 279/3944 (7.1%) |  |  |
| uMshwathi |  | 263/10304 (2.6%) | 166/6360 (2.6%) | 97/3944 (2.5%) |  |  |

<sup>1</sup>Incidence (%)
<sup>2</sup>Fisher's Exact Test
<sup>3</sup>Mean (SD)

There were 17,159 cases in wave 2 (Table 4) of which 59.7% were females and 37.7% were males, a further 437 cases had missing data on sex and were coded as missing values (NaN). The average age of patients in wave 2 was 41.55 years for females and 40.89 years for males (p=0.28).

**Table 4.** Descriptive analysis of SARS-CoV-2 cases, wave 2.

| Grouping Statistics by Gender | N | Overall, <sup>1</sup><br>N=17 159 | Female, <sup>1</sup><br>N=10 253 | Male, <sup>1</sup><br>N=6 469 | NaN, N=437 | P – value <sup>2</sup> | Incidence proportion<br>(Per 10 000 population) |
| --- | --- | --- | --- | --- | --- | --- | --- |
| Age | 16 926 | 41.31 (18.76)<sup>3</sup> | 41.55 (18.61)<sup>3</sup> | 40.89 (18.82)<sup>3</sup> | 42.20 (21.25)<sup>3</sup> | 0.28 |  |
| Age Group | 17 159 |  |  |  |  | <0.001 |  |
| 0-9 years |  | 2084/17159<br>(12.0%) | 1181/10253<br>(12.0%) | 847/6469<br>(13.0%) | 56/437<br>(13.0%) |  | 27.6 |
| 10-19 years |  | 1395/17159<br>(8.1%) | 821/10253<br>(8.0%) | 539/6469<br>(8.3%) | 35/437<br>(8.0%) |  | 62.5 |
| 20-29 years |  | 2691/17159<br>(16.0%) | 1677/10253<br>(16.0%) | 955/6469<br>(15.0%) | 59/437<br>(14.0%) |  | 122.1 |
| 30-39 years |  | 3357/17159<br>(20.0%) | 2051/10253<br>(20.0%) | 1241/6469<br>(19.0%) | 65/437<br>(15.0%) |  | 204.4 |
| 40-49 years |  | 3081/17159<br>(18.0%) | 1812/10253<br>(18.0%) | 1213/6469<br>(19.0%) | 56/437<br>(13.0%) |  | 279.3 |
| 50-59 years |  | 2245/17159<br>(13.0%) | 1327/10253<br>(13.0%) | 861/6469<br>(13.0%) | 57/437<br>(13.0%) |  | 345.5 |
| 60-69 years |  | 1401/17159<br>(8.2%) | 821/10253<br>(8.0%) | 520/6469<br>(8.0%) | 60/437<br>(14.0%) |  | 359.7 |
| 70-79 years |  | 672/17159<br>(3.9%) | 435/10253<br>(4.2%) | 221/6469<br>(3.4%) | 16/437<br>(3.7%) |  | 446.4 |
| 80+ years |  | 233/17159<br>(1.4%) | 128/10253<br>(1.2%) | 72/6469<br>(1.1%) | 33/437<br>(7.6%) |  | 487.2 |
| Municipality | 17 159 |  |  |  |  | <0.001 |  |
| Impendle |  | 534/17159<br>(3.1%) | 333/10253<br>(3.2%) | 147/6469<br>(2.3%) | 54/437<br>(12.0%) |  |  |
| Mkhambathini |  | 339/17159<br>(2.0%) | 206/10253<br>(2.0%) | 123/6469<br>(1.9%) | 10/437<br>(2.3%) |  |  |
| Mpofana |  | 522/17159<br>(3.0%) | 319/10253<br>(3.1%) | 180/6469<br>(2.8%) | 23/437<br>(5.3%) |  |  |
| Richmond |  | 1194/17159<br>(7.0%) | 767/10253<br>(7.5%) | 397/6469<br>(6.1%) | 30/437<br>(6.9%) |  |  |
| Msunduzi |  | 12764/17159<br>(74%) | 7554/10253<br>(74.0%) | 4915/6469<br>(76.0%) | 295/437<br>(68.0%) |  |  |
| uMngeni |  | 1042/17159<br>(6.1%) | 585/10253<br>(5.7%) | 446/6469<br>(6.9%) | 11/437<br>(2.5%) |  |  |
| uMshwathi |  | 764/17159<br>(4.5%) | 489/10253<br>(4.8%) | 261/6469<br>(4.0%) | 14/437<br>(3.2%) |  |  |

<sup>1</sup>Incidence (%)
<sup>2</sup>Fisher's Exact Test
<sup>3</sup>Mean (SD)

Msunduzi local municipality had the highest number of cases, 74.0% (p<0.001).

In wave 3 (Table 5), the aggregate number of cases was 20765, 56.5% were females, 42.8% were males, and 130 cases had missing data on sex and coded as missing values (NaN). Total cases in the age group 0-9 years increased to 14% of total cases compared to 12% in wave 2 and 3.4% in wave 1. For children in the age range 10-19 years, the total number of cases increased to 19% of total recorded cases compared to 8.1% in wave 2 and 6.9% in wave 1. Msunduzi local municipality contained the bulk of cases at 72%. Gendered differentiation across age and municipal differences were found to be statistically significant (p < 0.001).

**Table 5.**
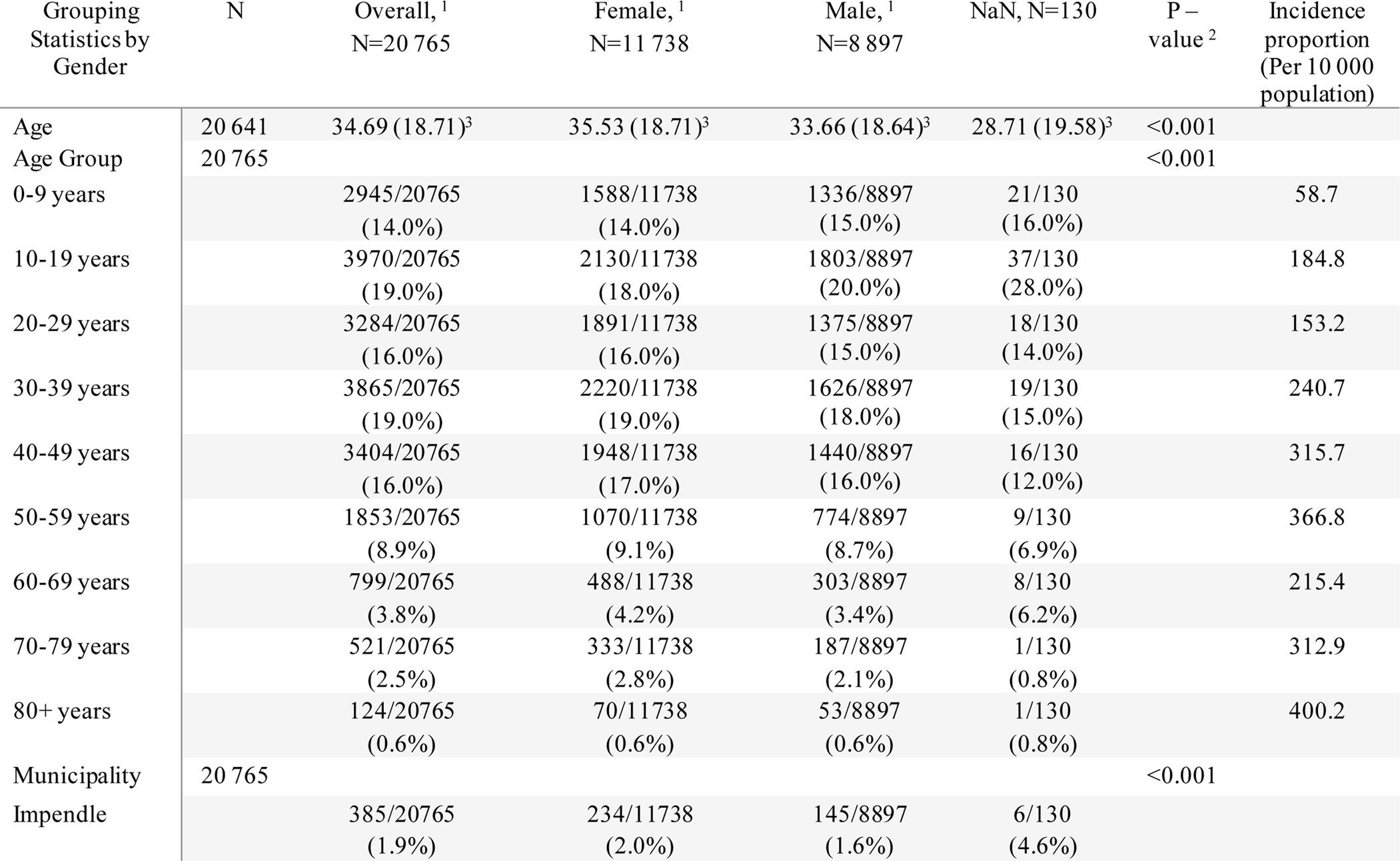
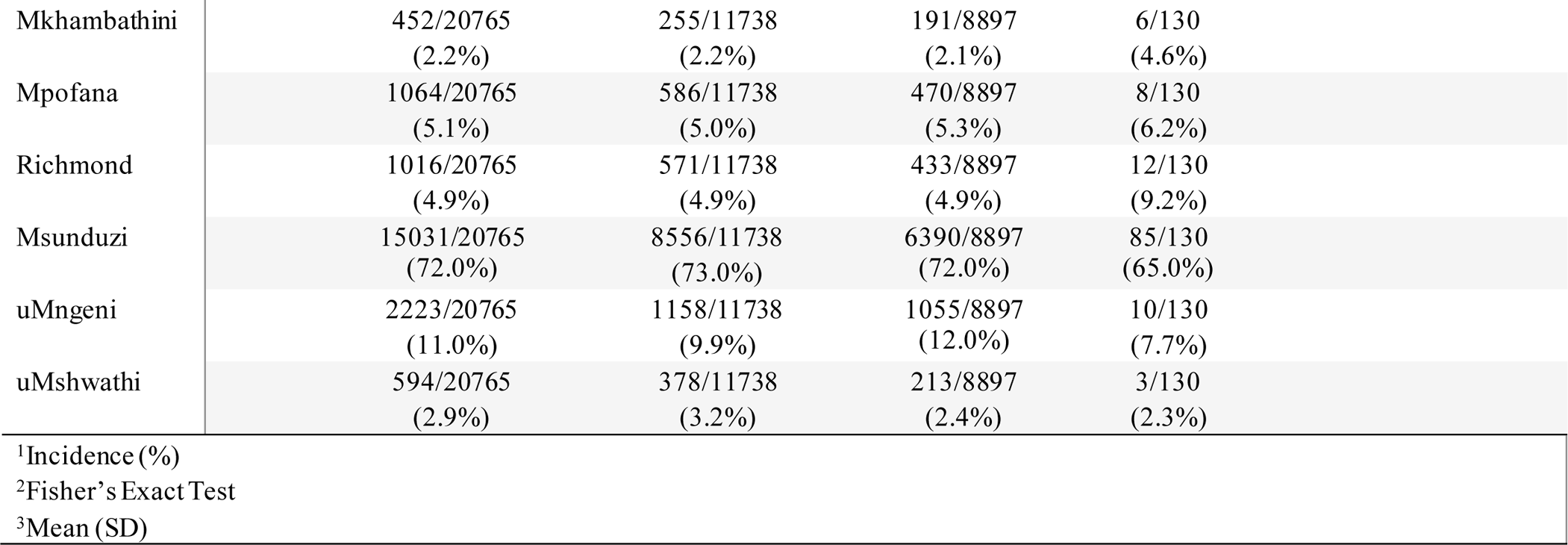
Descriptive analysis of SARS-CoV-2 cases, wave 3.

In wave 4, the total number of cases were 10569 as shown in Table 6. Of these reported cases, 57.9% were females, 39.9% were males, and 239 cases with missing gender data were coded NaN. Gendered differences in age were marginally statistically significant at the 5% level with a p-value of 0.058.

**Table 6.**
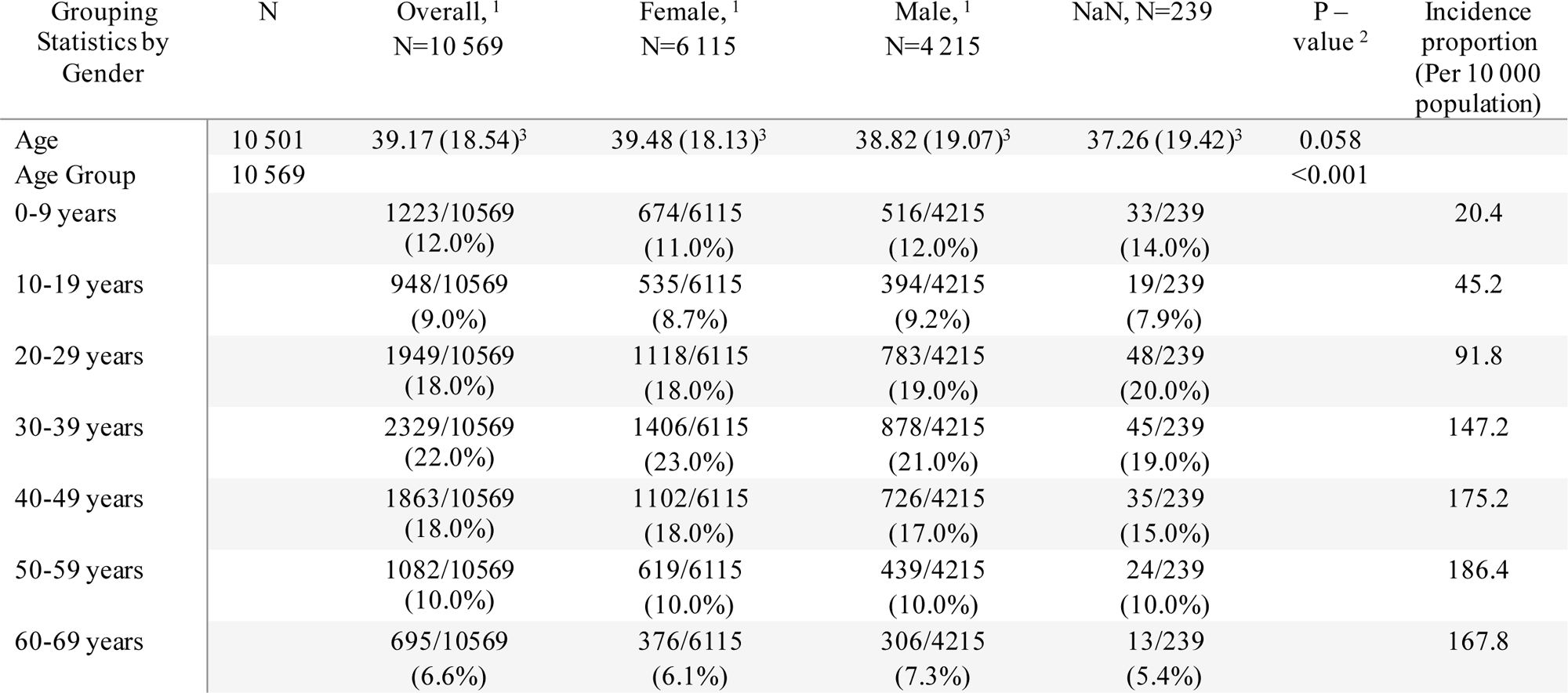
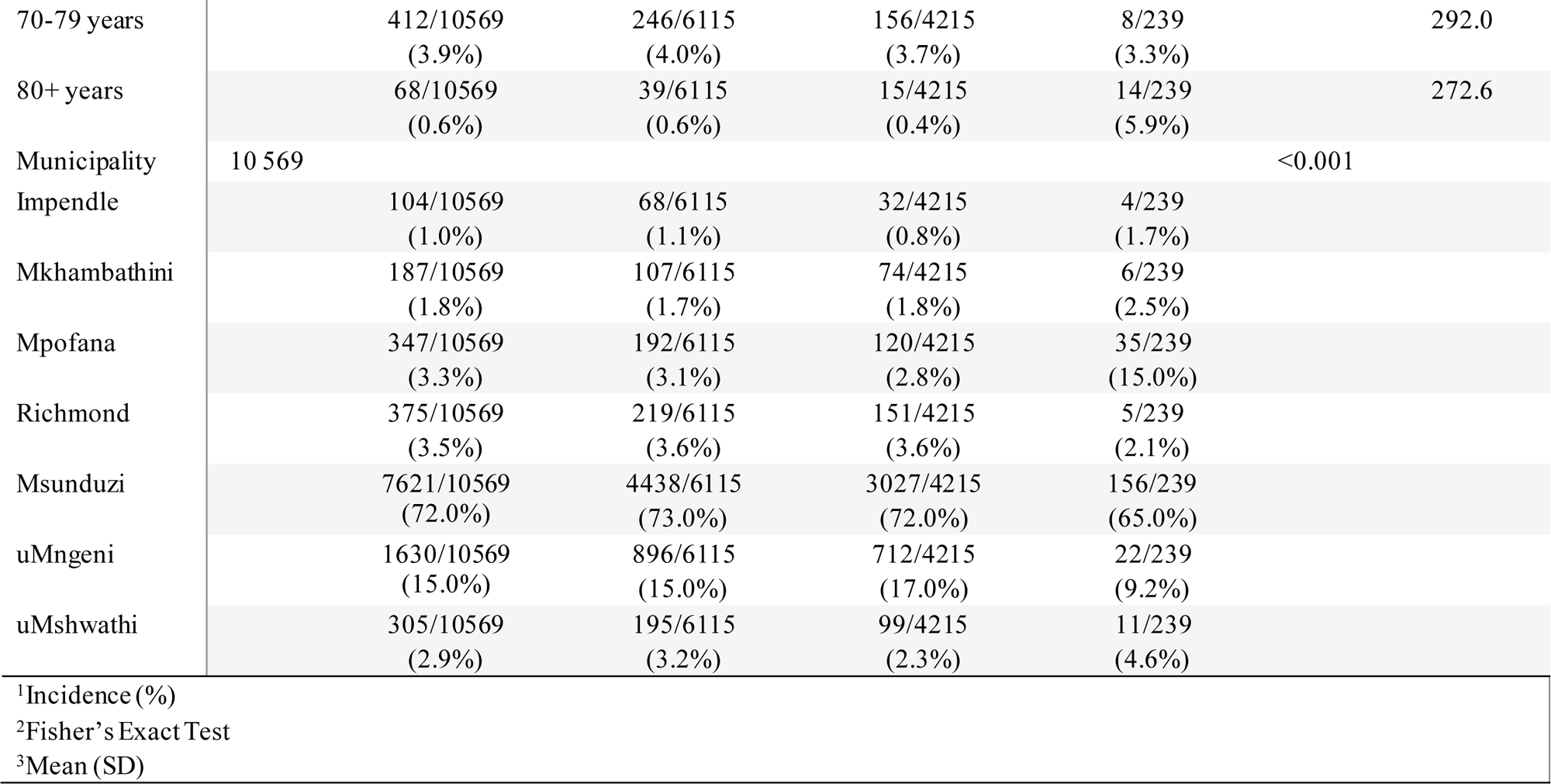
Descriptive analysis of SARS-CoV-2 cases, wave 4.

### Temporal analysis per wave of SARS-CoV-2 transmission

In wave 1 (Fig. 2), the time series plot shows incidence based on tests conducted from June 23, 2020, to August 30, 2020. Cases are seen to increase to a peak with a slightly more gradual reduction in cases towards the end of the period. From late June 2020, the number of positive cases were low, but a significant increase in positive cases was noted with the onset of July, which decreased towards the end of July. The largest positive cases were recorded during July, with large daily fluctuations observed. In South Africa, during the first wave, the predominant variant (>90% of viral sequences) was the ancestral strain with an Asp614Gly mutation (June to August 2020).

**Fig 2.**
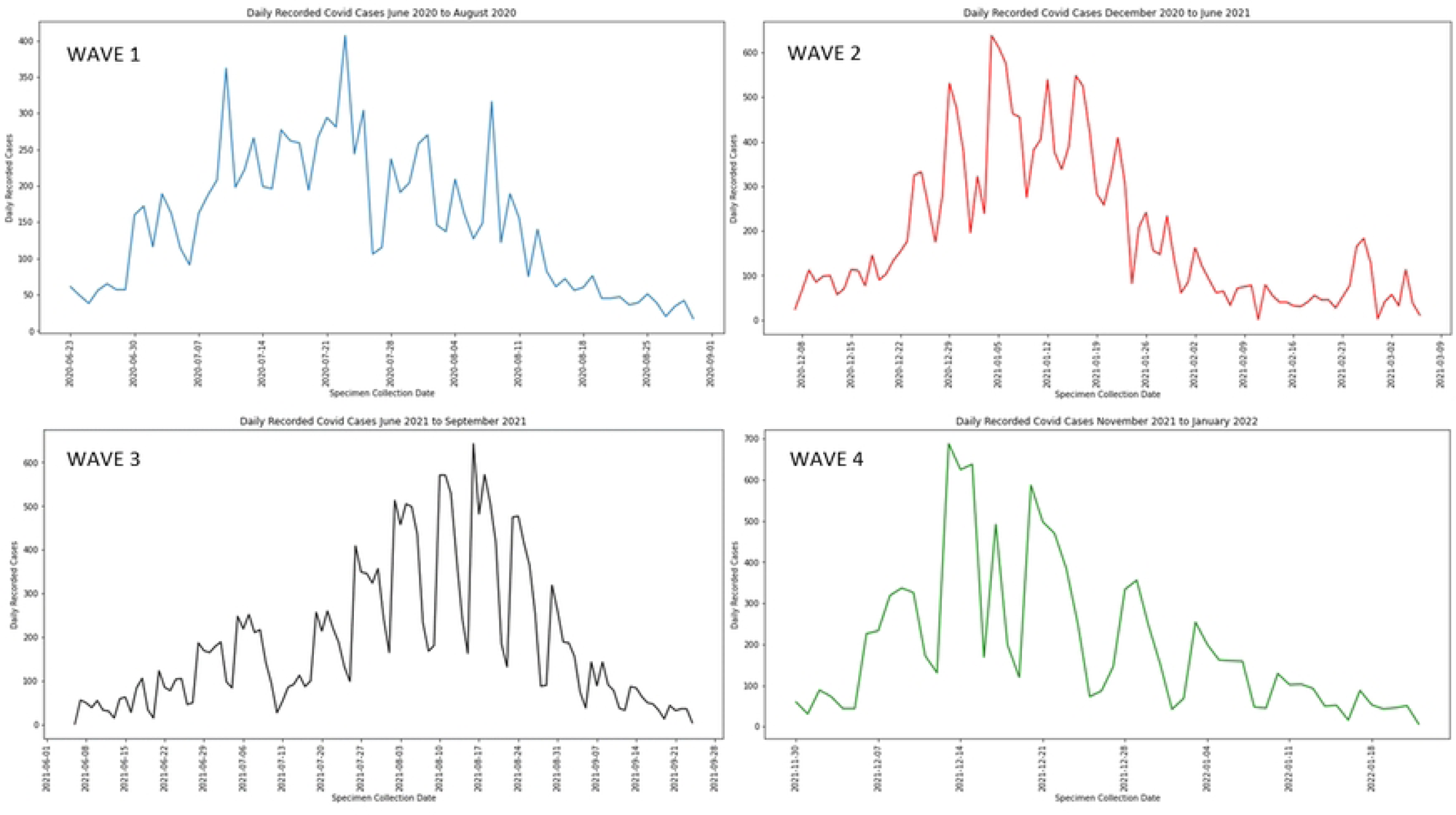
Time series plot of SARS-CoV-2 incidence, wave 1, uMgungundlovu District.

In wave 2 (Fig. 2), the time series plot shows incidence based on tests conducted from December 07, 2020, to March 06, 2021. During this wave, positive cases increase rapidly to a peak with a more gradual reduction towards the end of the period. Incidence can be observed to have fluctuated in the months between December 2020 and February 2021. A drop in the incidence is observed in mid-February, with fluctuations in trend decreasing as the wave transitions towards March 2021. Very high positive tests of up to 600 cases per day were recorded for the period between January 2021 and February 01, 2021. The predominant variant was the Beta variant.

In wave 3 (Fig. 2), the trend shows incidence for the period June 09, 2021, to September 25, 2021. There was a gradual increase in cases to a peak with a more rapid decline in cases towards the end of the period. Higher daily incidence is noted towards the middle of the period with daily fluctuation, within the beginning of the period, cases fluctuate at lower incidence. The end of the period shows less fluctuation with decreasing cases. In South Africa, during the third wave, the predominant variant was the Delta variant.

In wave 4 (Fig. 2), the trend shows incidence for the period November 30, 2021, to January 23, 2022. The wave is seen to peak early with large daily fluctuations and a more gradual decline in cases, fluctuations decrease towards the end of the period. The fourth wave is seen to be much shorter than the previous waves. In South Africa, during the fourth wave, Omicron was the predominant variant.

### Spatial analysis of SARS-CoV-2 transmission

Fig 3 illustrates the burden of transmission per sub-district relative to population size. Generally, the central portion of the district is affected by a higher incidence proportion, however, in wave 3 Mpofana district experienced a noticeable spike in cases.

**Fig 3.**
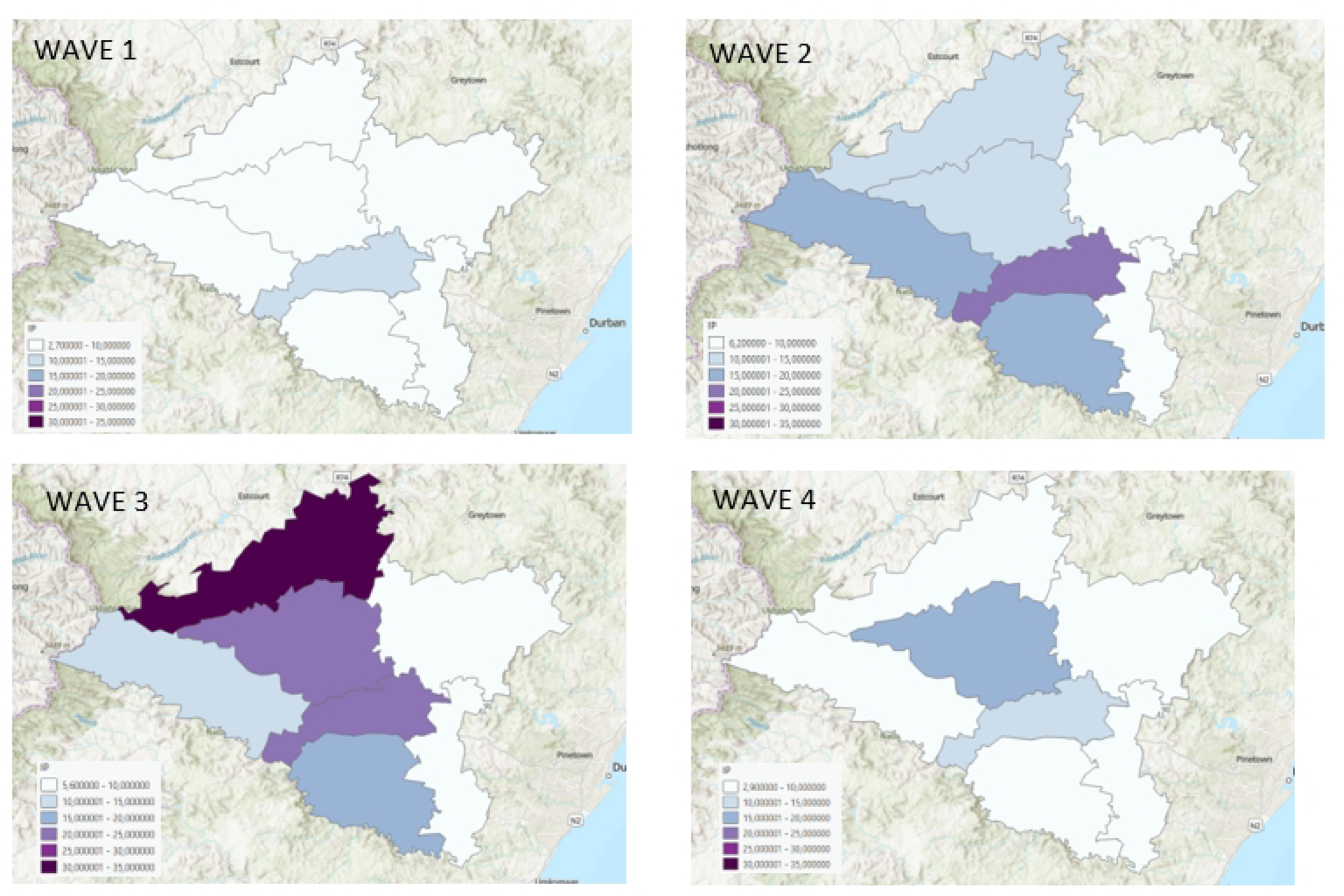
Choropleth map of SARS-CoV-2 incidence proportion per sub-district, per wave, uMgungundlovu district.

Fig 4 illustrates the burden of SARS-CoV-2 cases per ward. Within each sub-district, the wards that experienced higher transmission were the wards with higher economic activity. These included: Impendle ward 4; Richmond wards 1 and 2; uMngeni wards 2 and 6; Mkhambathini wards 1,3,4,5; Mpofana ward 1; Msunduzi wards 24, 25, 26, 27, 28, 29, 30, 31, 32, 33, 34, 35, 36 and uMshwathi wards 7 and 10.

**Fig 4.**
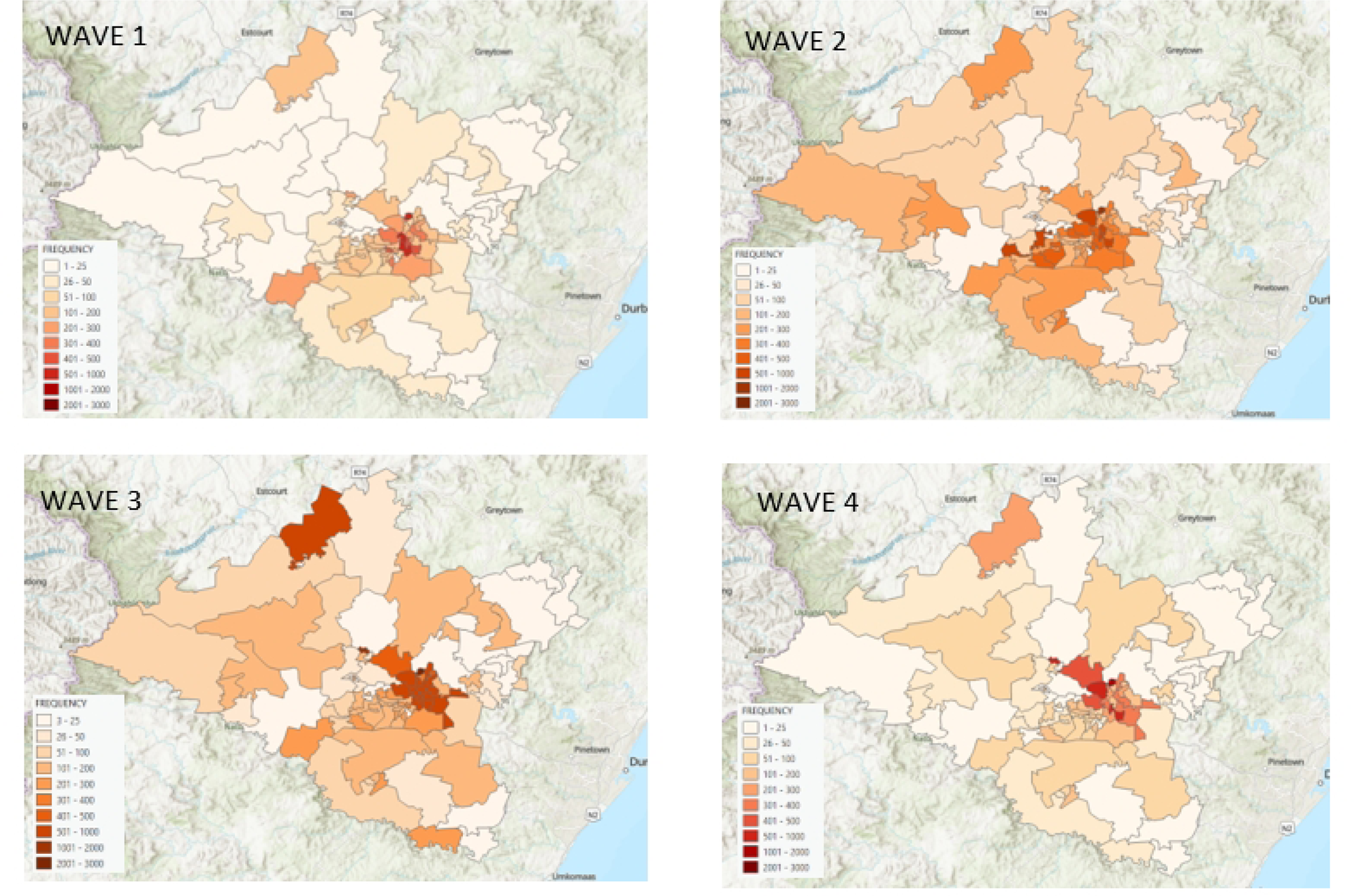
Choropleth map of SARS-CoV-2 incidence per ward, per wave, uMgungundlovu district.

### Kernel Density Estimation (KDE)

Areas of higher density per wave are nearly identical (Fig 5). Northern Msunduzi with the highest density of cases, followed by southern uMngeni, central Richmond, central Mpofana, southwestern Impendle, northern Mkhambathini and southern uMshwathi. The town areas of higher economic activity per sub-district are the areas with higher densities.

**Fig 5.**
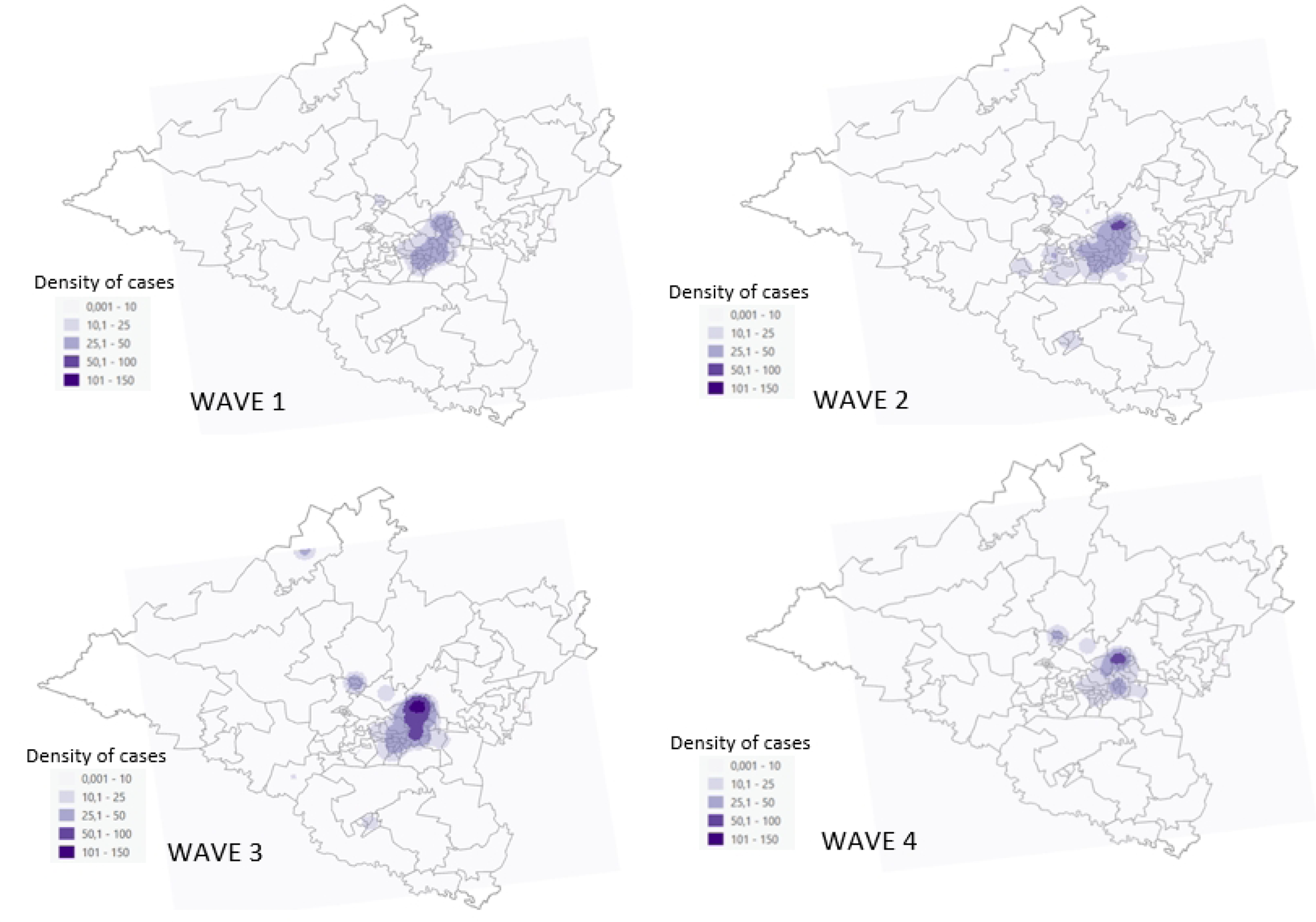
SARS-CoV-2 density maps per wave, uMgungundlovu district.

### Local indicators of spatial association (LISA)

During wave 1 (Fig 6) HH clusters were concentrated in the central wards of the district affecting northern Msunduzi and southern uMngeni. LL Clusters were found on the outskirts of the district affecting mostly uMshwathi, southern Richmond and Mkhambathini and a single ward in Impendle. In wave 2 HH clusters were concentrated in the central regions of the district affecting northern Msunduzi, northern Richmond and southern uMngeni. LL clusters were found on the outskirts of the district affecting most of uMshwathi, southern Richmond and Mkhambathini and a single ward in northern uMngeni. In wave 3 HH clusters were concentrated in the central regions of the district affecting northern Msunduzi, and southern uMngeni. LL clusters were found on the outskirts of the district affecting uMshwathi and Msunduzi. In wave 4 HH clusters were concentrated in the central regions of the district affecting northern Msunduzi, and southern uMngeni. LL clusters were found on the outskirts of the district affecting uMshwathi and Msunduzi, Richmond and Impendle.

**Fig 6.**
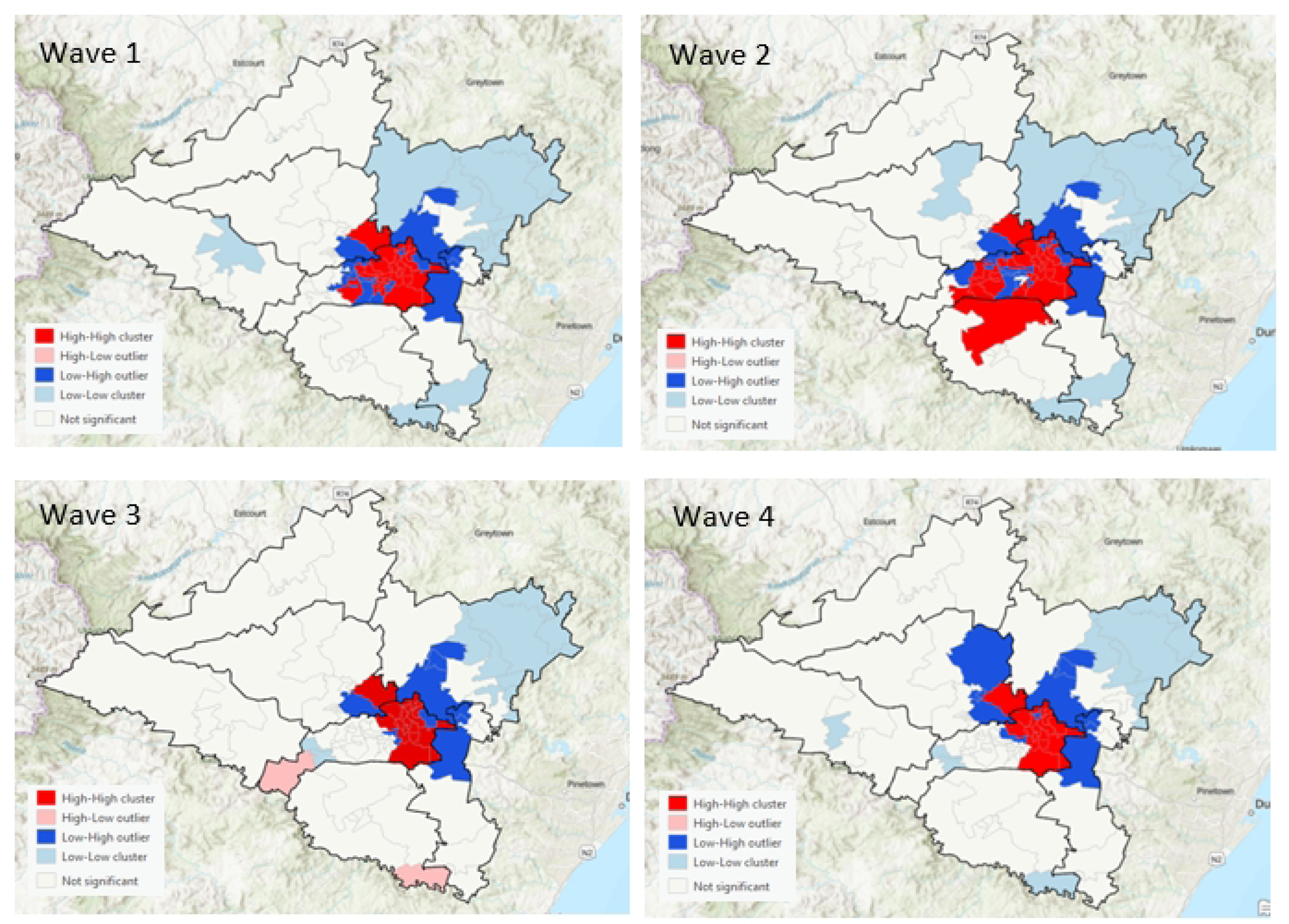
LISA analysis of SARS-CoV-2 incidence, uMgungundlovu district.

Positive spatial autocorrelation was found to occur during all waves (Moran’s I Wave 1=0.22; Wave 2=0.2; Wave 3=0.11; Wave 4=0.13).

In all waves, the data is skewed to the right, with more local municipalities having a lower frequency of cases and with fewer local municipalities having a high frequency of cases (wave 1: median=45.5; wave 2: median=141.5; wave 3: median=123; wave 4: median=55.5).

## Discussion

Across all four waves, females had the highest incidence of SARS-CoV-2. This could be due to higher health-seeking behaviour, work and relying on public transportation. Populations between the ages 20 and 50 experience the highest incidence, likely due to mobility for work, studies, and leisure. There is a clear spatial pattern of the spread of the virus within the district. The highest density of confirmed cases of COVID-19 occurred in the central town areas of each local municipality with a marked increase in the town areas of Msunduzi local municipality. In terms of spatial epidemiology, the built environment of higher economic activity and higher population density is highlighted as a strong contributor to increased transmission and a higher likelihood of contracting the virus. These are important indicators of priority areas and population groups to target when conducting public health interventions; similar findings were reflected in the literature.

### Demographic distribution of SARS-CoV-2 transmission

In the first wave, comparing incidence over age groups, it can be observed that the highest incidence was reported among the adult population within the age groups 20 to 29 years, 30 to 39 years, 40 to 49 years, and 50 to 59 years. These age groups are generally the most mobile due to work, school, and leisure. It may be reasonable to hypothesize that this could be a reason for increased transmission in these groups. It is also important to note that the population in question is a young population which may account for increased transmission in younger age groups, these findings are mirrored in a study conducted by de Souza et. al. (2021) on Brazilian populations with characteristics similar to the study population. Incidence proportion can be observed to increase associatively with age, which is attributed to the relatively young age cohort under analysis.

In the second wave, incidence among age groups changed remarkably, with the overall number of cases among children between 0-9 years rising to 12% compared to 3.4% in wave 1. In the third wave, the total cases among young children between the ages of 0-9 years rose to 14% of total cases from 12% in wave 2. For children in the age range 10-19 years, the total number of cases rose to 19% of total recorded cases in the third wave from 8.1% in wave 2. In wave 4 incidence continued to be higher in younger age categories than in wave 1. Before the 3^rd^ wave, vaccination efforts commenced within the district, however, the initial rollout of vaccines prioritised healthcare workers and high-risk population groups (the elderly and those living with underlying medical conditions) [14]. The increasing proportion of new infections in younger people has been equated to higher vaccination coverage in older population groups [15]. At the beginning of the pandemic, older and younger people engaged in similar preventive personal behaviours when controlling for other influences; however, as the pandemic progressed, older people adopted mitigating personal behavioural changes more than younger people [16]. Additionally, the variant of concern per wave could have contributed to the increase in paediatric cases.

This study documented a greater incidence of SARS-COV-2 infection among females compared to males. These findings, though contrary to earlier literature findings [23], are consistent with more recent research. Greater health-seeking behaviour associated with females when compared to males could account for these differences [7] and as such more women could have presented for testing than men resulting in a greater proportion of women testing positive. Within uMgungundlovu all symptomatic individuals presenting at public health facilities were tested, this could further explain the gender differences. Additionally, more women work in the informal sector and as essential workers, this may indicate that women occupy a higher percentage of occupations that may have put them at greater risk of contracting the virus [9].

### Spatial and Temporal Analysis

These findings add to a growing body of work examining population-level SARS-CoV-2 transmission, as well as differences in transmission across regions. The results of the first four waves observed have clearly illustrated that the central region of the district has been the most severely affected by SARS-CoV-2 transmission, this area is surrounded by local municipalities with significantly lower incidence for all waves except wave 3 during which Mpofana local municipality had a significant increase in SARS-CoV-2 incidence. This is interesting since according to the available research, regions adjacent to areas of high transmission will subsequently experience high transmission due to ‘spatial spillover’ [19]. Mkhambathini local municipality which borders eThekwini, a district which had the highest incidence of SARS-CoV-2 in the province, experienced relatively low incidence. Further research is required on possible explanatory or intervening factors beyond regional proximity.

Within uMgungundlovu District the first wave saw strict community level, household contact tracing and PCR testing of all close contacts, coupled with lockdown measures and fear of the unknown, most residents were willingly compliant. South Africa also experienced the first wave two months later than other countries thus benefitting from learning from other countries’ experiences [26]. These are likely contributing factors to the relatively less severe situation in the country and the district during the first wave compared to the global situation at the time [25]. During wave 1, high transmission was observed in Msunduzi local municipality, an economic hub of uMgungundlovu district known to be densely populated.

In the second wave of the pandemic, the infection rate was higher leading to an increase in the incidence of COVID-19 during that period [25]. The second wave spiralled when the National Corona Virus Command Centre shifted the lockdown to alert level 3 [31]. This allowed some degree of mobility in the country and among communities, and that interaction created a positive environment for rapid transmission of the virus in the district [26]. EThekwini district, adjacent to uMgungundlovu experienced the highest incidence of COVID-19 due to an increase in social activities that brought a multitude of people together. This increase in contact among community members spiraled the transmission of the virus. Additionally, incidence in the district of Harry Gwala which borders both Richmond and Impendle local municipalities is noted to increase approximately 2 weeks before incidence in Richmond and Impendle, this could be evidence of ‘spatial spill-over’ [20].

Transmission of SARS-CoV-2 and the resulting case load was seen to be much higher in the 3rd wave. This among others, could have been due to the social unrest that occurred in the district due to the continued lockdown. The widespread unrest that led to massive damage to property and infrastructure was catalytic to the widespread transmission of the virus in the area [28]. It is believed that this social upheaval that undermined public health preventive measures was a result of lockdown fatigue, poor social distancing, and slow vaccine rollouts. [27]. Mpofana sub-district experienced the highest incidence proportion. This may be due to a relaxation of restrictions involving international trade and an increase in congregate setting cases.

During wave 4, transmission of the Corona Virus looked different from the first three waves, in that the wave was shorter, and the cases spiked quicker reaching a peak within a much shorter time as opposed to what was observed with the initial 3 waves. This could have been due to other reasons such as vaccination, which was extensively underway during this time.

The LISA analysis undertaken showed that the distribution of incidence of SARS-CoV-2 in uMgungundlovu was clustered. An increase in SARS-CoV-2 infection was significantly higher in the HH wards which were areas of higher economic activity, population density and the built environment. Wards with statistically significant LL clustering remained on the outskirts of the district, in rural areas.

The Kernel density estimation showed that areas with the highest density of confirmed SARS-CoV-2 cases occurred in the central town areas of Msunduzi sub-district and, to a lesser extent, in the central town areas of the rest of the local municipalities. Clearly, lower densities were noted in rural areas. This might have been an indication of increased contact within areas of higher economic activity as well as population density hence contributing to increased transmission. This may also be an indication of reduced access to testing facilities in rural areas hence an underestimation of the actual burden of the COVID-19 pandemic.

## Conclusion

Our study appears to be the only one to-date to document the spatial distribution of SARS-CoV-2 in uMgungundlovu District, KwaZulu Natal, South Africa. The findings emanating from this study consolidate the epidemiological understanding of the main key drives of SARS-COV-2 infection in this diverse population. The observed higher concentration of case distributions observed in the northern regions of Msunduzi, might show a clear relationship of infection transmission and increased economic activity, population density and urban development. The data from this study shows that transmission rates across the district were not uniform and were driven by increased community mixing in urban areas. Further research could help expose further, the dynamics of urban living and infection disease epidemics and explore feasible and easy to implement public health interventions that could curtail a rising epidemic with existing health system structures.

## Data Availability

The data underlying the results presented in the study are available from the Kwa-Zulu Natal Department of Health, uMgungundlovu District.

https://dataportal-mdb-sa.opendata.arcgis.com/maps/97cb14748cef423a94f7b7154387b2dc

## Acknowledgements

Radiya Gangat, a postgraduate student in Epidemiology was financially supported by the Stellenbosch University. The fellowship facilitated completion of the research project from which this manuscript was derived. We thank the National Institute for Communicable Diseases and the uMgungundlovu Health District Office for their support of this project.

